# Emergency Department Utilization Following a Community Hospital Closure: A Population-Based Comparative Study

**DOI:** 10.64898/2026.10.21.26366109

**Authors:** Simon Paul

**Affiliations:** Madera County Department of Public Health Madera, California, USA

## Abstract

**Importance:** Hospital closures may alter access to emergency care. However, changes in utilization among residents of affected communities are less well characterized than changes in volume at surrounding hospitals.

**Objective:** To compare changes in emergency department (ED) utilization among Madera County residents following the closure of Madera Community Hospital (MCH) with changes among Fresno County residents, and to examine geographic differences by pre-closure reliance on MCH.

**Design, Setting, and Participants:** Retrospective population-based comparative study of annual California hospital Patient Origin/Market Share data, 2015-2024, with an exploratory comparative interrupted time-series analysis. Pre-pandemic years were 2015-2019, pandemic-affected pre-closure years were 2020-2022, and the two complete closure years were 2023-2024. Encounters were classified by county of patient residence, regardless of treating hospital location.

**Main Outcomes and Measures:** Combined ED encounters (ED-only plus inpatient-from-ED) and each component separately. County-level utilization was expressed as encounters per 1,000 residents using annual population estimates. Geographic comparisons by pre-closure MCH dependence used ZIP-code encounter counts.

**Results:** From 2022 to 2023, the first complete year following closure, combined ED utilization declined from 398.47 to 333.61 encounters per 1,000 Madera County residents (−16.3%), compared with 399.61 to 398.49 (−0.3%) among Fresno County residents. Madera ED-only rates declined 20.4%, while inpatient-from-ED rates increased 13.4%; however, the adjusted comparative analysis did not identify a consistent additional Madera-specific increase in inpatient-from-ED utilization. Exploratory regression estimated Madera-specific closure-period rate ratios of 0.871 for combined ED utilization and 0.848 for ED-only utilization. Combined ED encounter counts declined 18.4% in high-MCH-dependence ZIP codes versus 5.8% in low-dependence ZIP codes. Statistical inference was limited by the inclusion of only two counties, two complete closure years, and substantial overdispersion.

**Conclusions and Relevance:** The period following MCH closure was associated with a substantial reduction in ED utilization among Madera County residents relative to Fresno County residents, concentrated in ED-only encounters and communities previously most dependent on MCH. The observational design does not establish causation or determine whether reduced utilization reflected substitution to other care settings, decreased demand, or unmet medical need.

## Introduction

Hospital closures are an increasing concern in the United States, particularly among smaller hospitals and those serving rural or financially vulnerable communities. As of July 2026, approximately 700 rural hospitals nationwide were estimated to be at risk of closure, including 264 at immediate risk.^1^ In California, four hospitals had closed or narrowly avoided closure during the preceding three years, and additional hospitals had discontinued clinical service lines.^2^ Understanding the consequences of hospital closures is important for affected communities and for planning emergency medical services, regional hospital capacity, and access to care in communities where hospitals remain vulnerable.

Hospital closures may have adverse economic consequences for communities and alter access to medical care, particularly when residents must travel farther to obtain emergency services. Previous studies have documented a range of clinical and utilization outcomes, with consequences potentially dependent on the availability and proximity of alternative facilities.^3,4^ Studies of hospitals receiving patients after nearby closures have reported increased emergency department (ED) and inpatient workloads.^5^ However, increases in utilization at receiving hospitals do not necessarily establish that residents of the community experiencing the closure have maintained their previous level of access to care. Indeed, one study of commercially insured residents affected by rural hospital closures found reductions in both ED and outpatient utilization.^6^ Assessing utilization by patient residence, rather than solely by destination facility, provides a complementary perspective on the consequences of hospital closure.

Madera Community Hospital (MCH) was the only general acute care hospital serving adults in Madera County, California, a geographically diverse county encompassing urbanized communities, agricultural areas, and rural mountain communities. Its ED closed on December 30, 2022, with remaining hospital services ending January 3, 2023. The hospital reopened on March 18, 2025.^7,8^ Before closure, Madera County residents received emergency care both at MCH and at hospitals in neighboring Fresno County, with substantial variation in reliance on MCH across residential ZIP codes. Following closure, the nearest alternative general adult emergency department was approximately 23 miles away in Fresno County, although the additional travel distance varied according to patients’ locations of residence.

California’s publicly available hospital utilization datasets permit examination of ED encounters by patient county and ZIP code of residence, as well as by destination hospital, providing an opportunity to assess changes in utilization among the population affected by the closure.^9^ ED utilization is particularly relevant because emergency encounters encompass a wide range of clinical conditions and levels of acuity. A decline in total ED encounters among residents of a community following a hospital closure could reflect several mechanisms, including substitution to outpatient settings, reductions in discretionary visits, or forgone care. Increased travel distances, transportation difficulties, and financial constraints may contribute to these changes, potentially affecting patients differently according to clinical urgency and socioeconomic resources. Distinguishing ED-only encounters from those resulting in inpatient admission may provide additional insight into changes in encounter composition, although admission status is not a direct measure of clinical severity.

We examined annual population-adjusted ED utilization among Madera and Fresno County residents from 2015 through 2024, encompassing the period before closure and the two complete calendar years following closure. We compared ED-only and inpatient-from-ED encounters, examined changes in hospital destinations, and assessed whether utilization changes were concentrated in Madera County ZIP codes with greater historical dependence on MCH. Fresno County residents and Madera County communities with relatively low pre-closure reliance on MCH provided complementary geographic comparisons. Because the COVID-19 pandemic substantially disrupted hospital utilization during the study period, comparative interrupted time-series methods were used to evaluate the association between the closure period and Madera County ED utilization while accounting for underlying trends and pandemic-period changes.^10,11^

## Methods

### Study design and setting

This retrospective ecological study examined annual ED utilization among Madera and Fresno County residents before and after the closure of MCH. MCH’s ED closed on December 30, 2022. The study included 2015–2024, with 2015–2019 representing the pre-pandemic period, 2020–2022 the pandemic-affected pre-closure period, and 2023–2024 the two complete calendar years following closure. Calendar year 2025 was excluded because MCH reopened in March of that year. Fresno County residents served as an external comparison population because of geographic proximity, broadly similar pre-closure ED utilization rates and trends, and shared regional healthcare resources. Fresno County was not assumed to be unaffected by MCH closure, since its hospitals also received patients from Madera County.

### Data sources and outcome definitions

Annual ED encounter data were obtained from the California Department of Health Care Access and Information (HCAI) Patient Origin/Market Share dataset.^9^ Encounters were classified according to the patient’s reported county or ZIP code of residence rather than the location of the treating hospital. The dataset includes encounters at reporting California hospitals, regardless of destination county. For the county-level analysis, records not assigned to Madera or Fresno County were excluded; unknown residence was not imputed or reassigned.

Two encounter categories were analyzed separately. ED Only represents ED treat-and-release encounters that did not result in inpatient admission at the same hospital. Inpatient from ED represents inpatient discharges originating in the hospital’s ED; these admissions are included in the inpatient data rather than counted as separate ED-only encounters. Combined ED encounters were defined as the sum of ED Only and Inpatient from ED.

Annual encounter counts represent encounters rather than unique patients or linked episodes of care. Repeat visits and care at multiple hospitals may contribute more than one encounter.

Additional HCAI county-of-residence ED characteristics datasets were used to describe ED-only treat-and-release encounters by expected payer, race/ethnicity, and sex in 2022 and 2023.^12^ These datasets exclude encounters resulting in same-hospital inpatient admission and were therefore analyzed separately from combined ED utilization. Expected payer categories were grouped into uninsured/self-pay, Medi-Cal, Medicare, private insurance, and other. Subgroup comparisons used encounter counts rather than payer- or demographic-specific population-adjusted rates.

Hospital destinations for Madera County residents were examined using the Patient Origin/Market Share data. Combined ED encounters were grouped according to the county of the treating hospital, using the same encounter definition as the primary county-level analysis.

### Population denominators and data validation

Annual county ED utilization rates were calculated by dividing encounter counts by the corresponding July 1 county population estimate and multiplying by 1,000. Population denominators were obtained from California Department of Finance E-2 county population estimates, using the historical 2010–2019 series for study years 2015–2019 and the February 2026 revised 2020–2025 series for study years 2020–2024.^13,14^

Source data were downloaded and processed using reproducible Python scripts. Aggregated encounter counts and calculated ED utilization rates were checked against the original source data, and all final county rates were independently reproduced from the validated encounter counts and population denominators.

### Geographic comparison within Madera County

Because Madera County includes communities with substantially different geographic access to hospitals, a secondary analysis examined whether changes in ED utilization differed according to residents’ historical reliance on MCH.

For each Madera County residential ZIP code, MCH dependence was calculated as the percentage of combined ED encounters occurring at MCH among all combined ED encounters by residents of that ZIP code during 2017–2022. Based on observed gaps in these pre-closure percentages, ZIP codes were classified into three groups:

- **High dependence:** 93638, 93639, 93637, and 93610 (approximately 47%–57% of combined ED encounters at MCH).
- **Intermediate dependence:** 93653 and 93636 (approximately 16%–29%).
- **Low dependence:** 93669, 93643, 93644, 93645, 93614, 93601, and 93604 (approximately 3%–7%).

The ZIP-code level comparison examined high- and low-dependence areas. Intermediate-dependence ZIP codes were excluded from this comparison to provide a clearer contrast between communities with substantially different historical reliance on MCH.

These classifications were based exclusively on pre-closure ED utilization and were not determined using post-closure changes. The high-dependence group was concentrated primarily along the Highway 99 corridor, while low-dependence areas included communities more oriented toward Fresno-area hospitals, including those along Highway 41.

ZIP-code group analyses compared annual encounter counts and percentage changes rather than population-adjusted rates because validated annual population denominators were not available for every included ZIP code.

### Statistical analysis

The primary analysis was descriptive. Annual ED utilization rates were compared between Madera and Fresno counties, with particular attention to changes from 2022 to 2023 and 2024. Absolute and percentage changes were calculated for combined ED encounters, ED-only encounters, and inpatient-from-ED encounters. Descriptive trends and annual rates of change during the pre-pandemic period (2015–2019) were examined separately from pandemic-affected years (2020–2022).

As a secondary exploratory analysis, comparative interrupted time-series regression was used to assess whether differences between Madera and Fresno persisted after accounting for pre-existing trends and pandemic-period changes.

Poisson regression models were fitted to annual encounter counts, using the natural logarithm of annual county population as an offset to account for differences in population size. The models allowed the counties to have different baseline utilization rates, different underlying time trends, and different changes during the pandemic period. They also included a term representing the closure years (2023–2024) and an interaction between Madera County and the closure period.

The Madera-by-closure interaction estimated the additional relative change in Madera’s ED utilization rate during 2023–2024 compared with Fresno, after adjustment for the model’s other terms. Results were expressed as rate ratios. These estimates were interpreted as associations with the closure period rather than definitive causal effects of MCH closure.

Sensitivity analyses examined alternative assumptions about pre-closure trends, exclusion of pandemic-affected years, and separate estimates for 2023 and 2024.

Because the regression analysis included only two counties and two complete post-closure years, and because the count models exhibited substantial overdispersion, the model results were interpreted cautiously. Emphasis was placed on the observed descriptive patterns and consistency of model estimates across alternative specifications rather than formal statistical significance.

All data processing and statistical analyses were performed using locally executed Python scripts. Encounter data were queried using DuckDB and processed using pandas and NumPy. Descriptive trend analyses used SciPy, while Poisson regression models were fitted using statsmodels. Figures were generated using Matplotlib. The principal statistical models were independently refitted using a separate Python validation script, confirming reproducibility of the reported estimates.

### Use of Generative Artificial Intelligence

Generative artificial intelligence (ChatGPT, OpenAI) was used to assist with identifying public data sources, developing the analytical approach, generating and refining Python code, interpreting preliminary results, and drafting and editing the manuscript. All source data were obtained and processed locally, and statistical analyses were performed using locally executed Python scripts. Numerical results were independently validated against source data and reproducible analytical outputs. The author reviewed and verified the analyses, independently evaluated the interpretations, revised the manuscript, and assumes full responsibility for its content.

### Ethics and reporting

The study protocol was reviewed by the Fresno County Institutional Review Board and determined to be exempt from IRB review on October 13, 2026.

## Data availability

All source data used in this study are publicly available from the California Department of Health Care Access and Information and the California Department of Finance through the websites cited in the References. No restricted patient-level data were used.

## Results

### Baseline utilization and pre-closure patterns

During 2017–2019, Madera County residents (mean population, 155,002) averaged 66,417 combined ED encounters annually, compared with 436,184 encounters among Fresno County residents (mean population, 993,666). Corresponding averages of the annual utilization rates were 428.52 and 439.04 encounters per 1,000 residents, respectively. The ED-only rates were 380.24 and 384.79, and inpatient-from-ED rates were 48.29 and 54.25, respectively. Both counties’ combined ED rates declined before the pandemic, with estimated annual changes of −1.85% for Madera and −1.30% for Fresno during 2015–2019. The pre-pandemic slopes were similar in magnitude but not identical, and the comparison does not establish parallel trends. Both series fell sharply in 2020 and recovered unevenly through 2022 (Figure 1).

**Figure 1.**
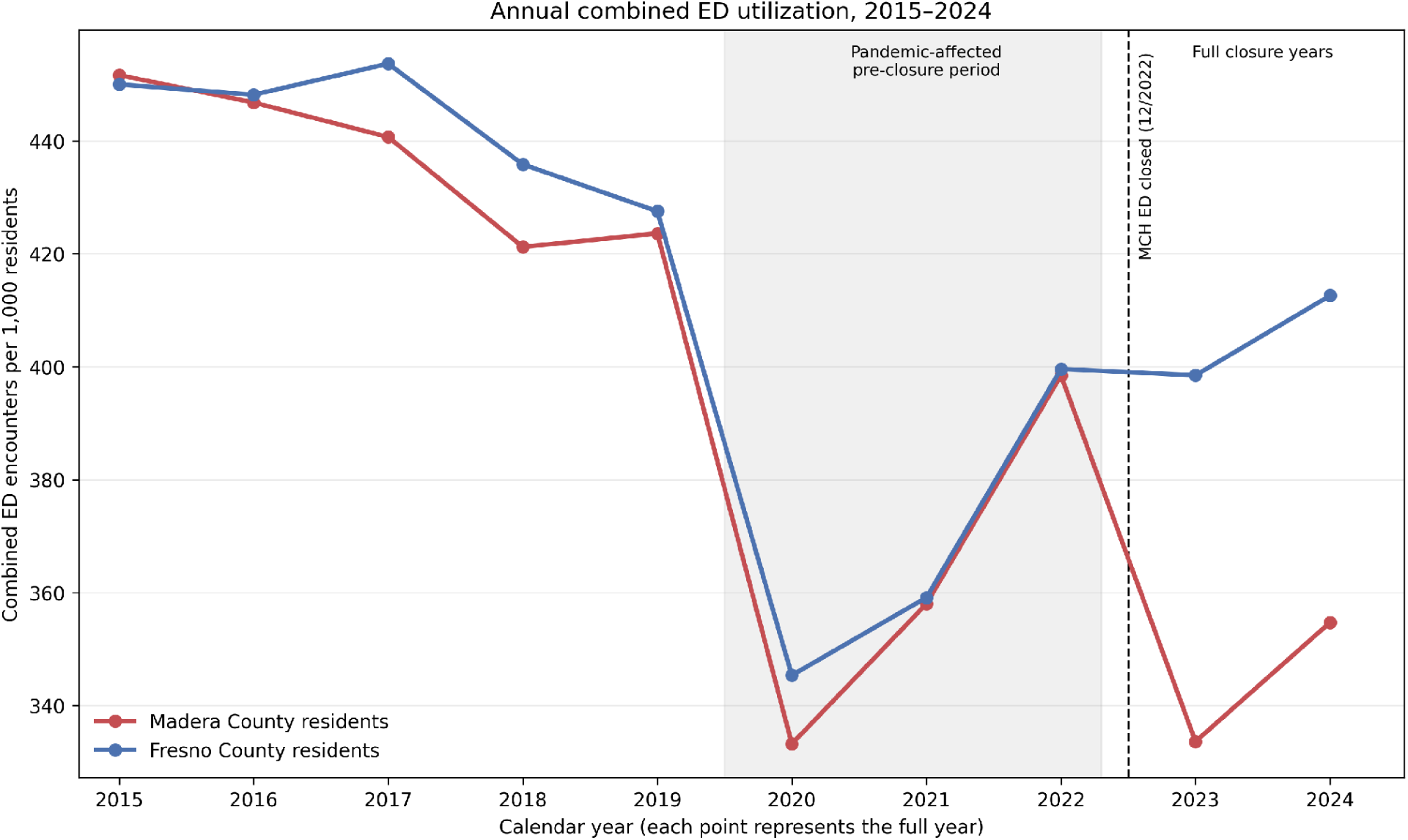
Annual combined emergency department (ED) encounters per 1,000 Madera and Fresno County residents, 2015–2024. Rates were calculated using annual county population estimates. The shaded area identifies the pandemic-affected pre-closure period (2020–2022), and the vertical dashed line marks the closure of the Madera Community Hospital (MCH) emergency department on December 30, 2022.

### County-level changes after closure

In 2022, combined ED rates were 398.47 per 1,000 Madera residents and 399.61 per 1,000 Fresno residents. Following the December 2022 closure of MCH’s emergency department, the 2023 rates fell to 333.61 and 398.49, respectively: absolute changes of −64.87 and −1.12 per 1,000, corresponding to −16.28% and −0.28% (Table 1; Figure 1). The Madera rate partially recovered to 354.71 in 2024, remaining 10.98% below 2022, while Fresno increased to 412.61 (3.25% above 2022). Relative to 2017–2019 mean rates, the 2023–2024 mean was 19.69% lower in Madera and 7.63% lower in Fresno.

**Table 1.**
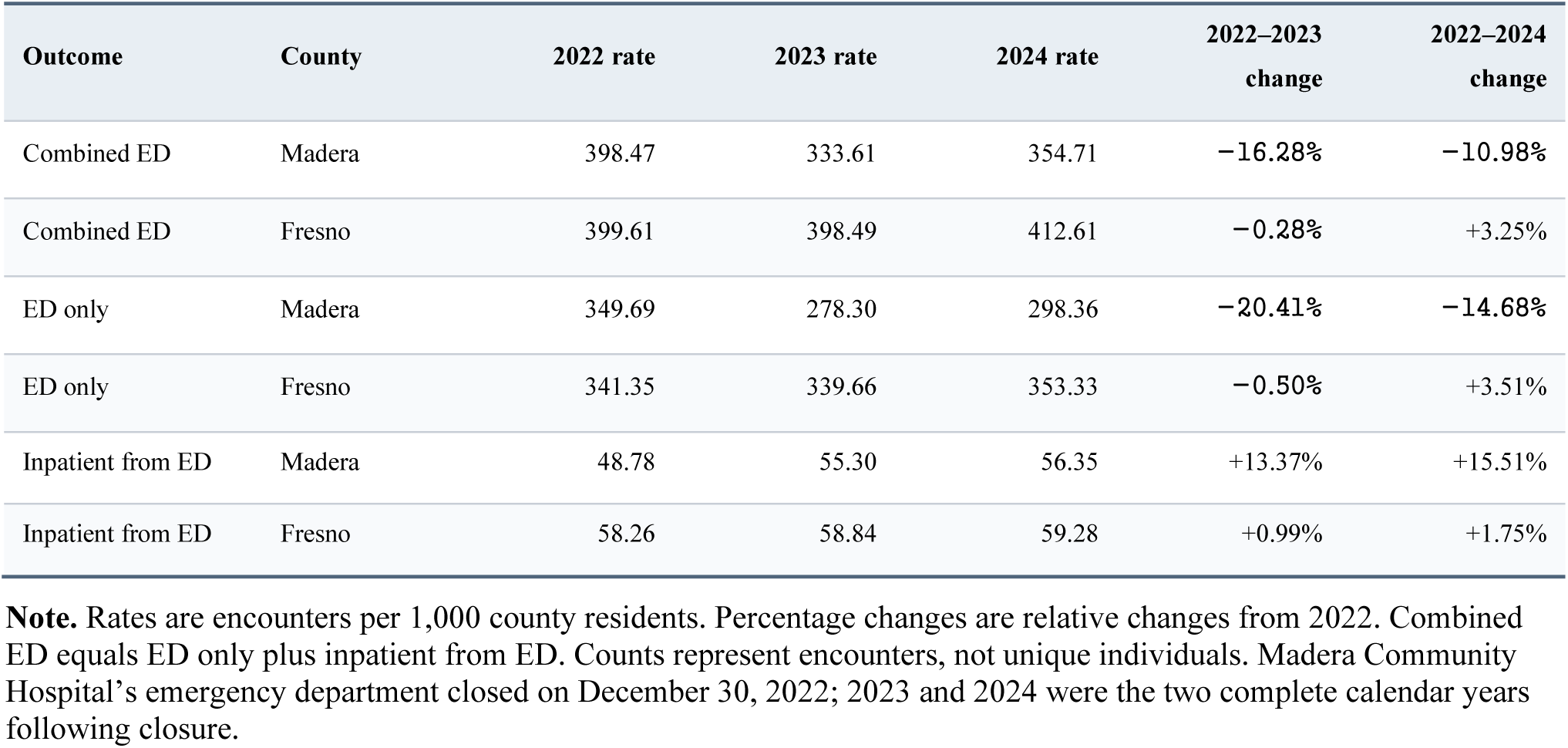
County-level emergency department utilization rates and changes, 2022–2024.

The decline in utilization in Madera was concentrated in ED-only encounters (Figure 2). From 2022 to 2023, Madera ED-only rates declined from 349.69 to 278.30 per 1,000 residents (−20.41%), compared with 341.35 to 339.66 in Fresno (−0.50%). In contrast, Madera inpatient-from-ED rates increased from 48.78 to 55.30 (+13.37%), while Fresno increased from 58.26 to 58.84 (+0.99%). In 2024, the pattern persisted: Madera’s ED-only rate remained 14.68% below its 2022 level, while its inpatient-from-ED rate was 15.51% higher. However, the comparative regression analysis did not identify a consistent additional increase in inpatient-from-ED rates specific to Madera after accounting for prior trends and pandemic-period changes.

**Figure 2.**
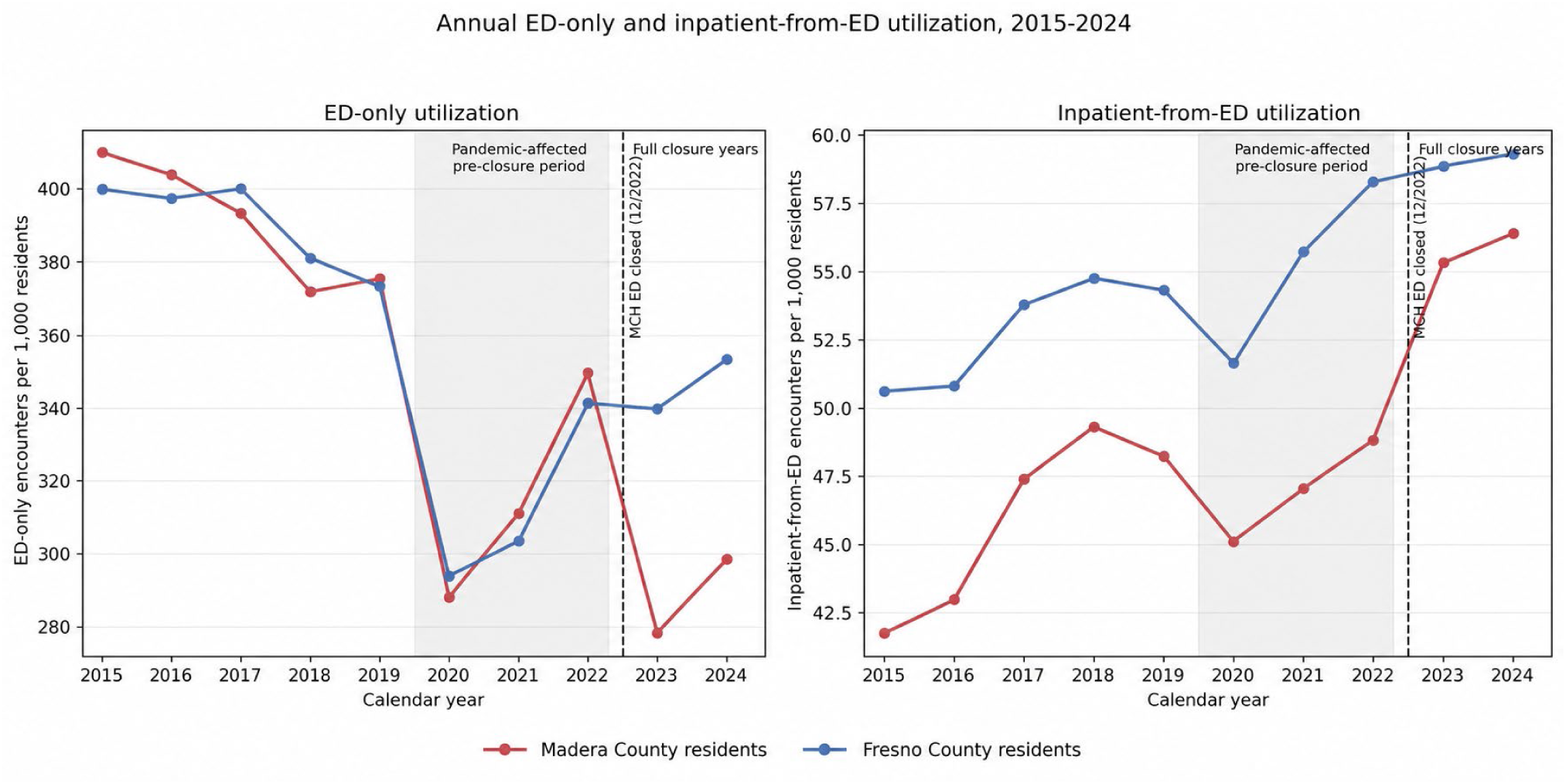
Annual ED-only and inpatient-from-ED encounters per 1,000 Madera and Fresno County residents, 2015–2024. ED-only encounters are shown in the left panel and inpatient-from-ED encounters in the right panel. The panels use different vertical scales. The shaded area identifies the pandemic-affected pre-closure period (2020–2022), and the vertical dashed line marks the closure of the Madera Community Hospital (MCH) emergency department on December 30, 2022.

### Geographic variation in utilization

Among Madera County ZIP codes previously highly dependent on MCH (93638, 93639, 93637, and 93610), combined ED encounter counts declined from 50,497 in 2022 to 41,202 in 2023 (−18.4%). The seven low-dependence ZIP codes experienced a smaller decline, from 7,712 to 7,266 encounters (−5.8%). The difference in percentage changes between the groups was approximately 12.6 percentage points. In 2024, encounter counts in the high-dependence group remained 12.1% below their 2022 level, compared with 1.5% below baseline in the low-dependence group (Table 2; Figure 3).

**Table 2.** Combined ED encounter counts by pre-closure MCH dependence among Madera County ZIP-code groups.

| MCH dependence | 2022 encounters | 2023 encounters | 2024 encounters | 2022–2023 change | 2022–2024 change |
| --- | --- | --- | --- | --- | --- |
| High (4 ZIPs) | 50,497 | 41,202 | 44,398 | –18.4% | –12.1% |
| Low (7 ZIPs) | 7,712 | 7,266 | 7,599 | –5.8% | –1.5% |

**Note.** High-dependence ZIP codes: 93638, 93639, 93637, 93610. Low-dependence ZIP codes: 93669, 93643, 93644, 93645, 93614, 93601, 93604. Groups were classified by pre-closure MCH encounter shares; intermediate ZIP codes 93636 and 93653 were excluded from this comparison. Combined ED includes ED-only and inpatient-from-ED encounters among residents at reporting California hospitals, regardless of destination. Counts are encounters, not unique patients, and are not population-adjusted.

**Figure 3.**
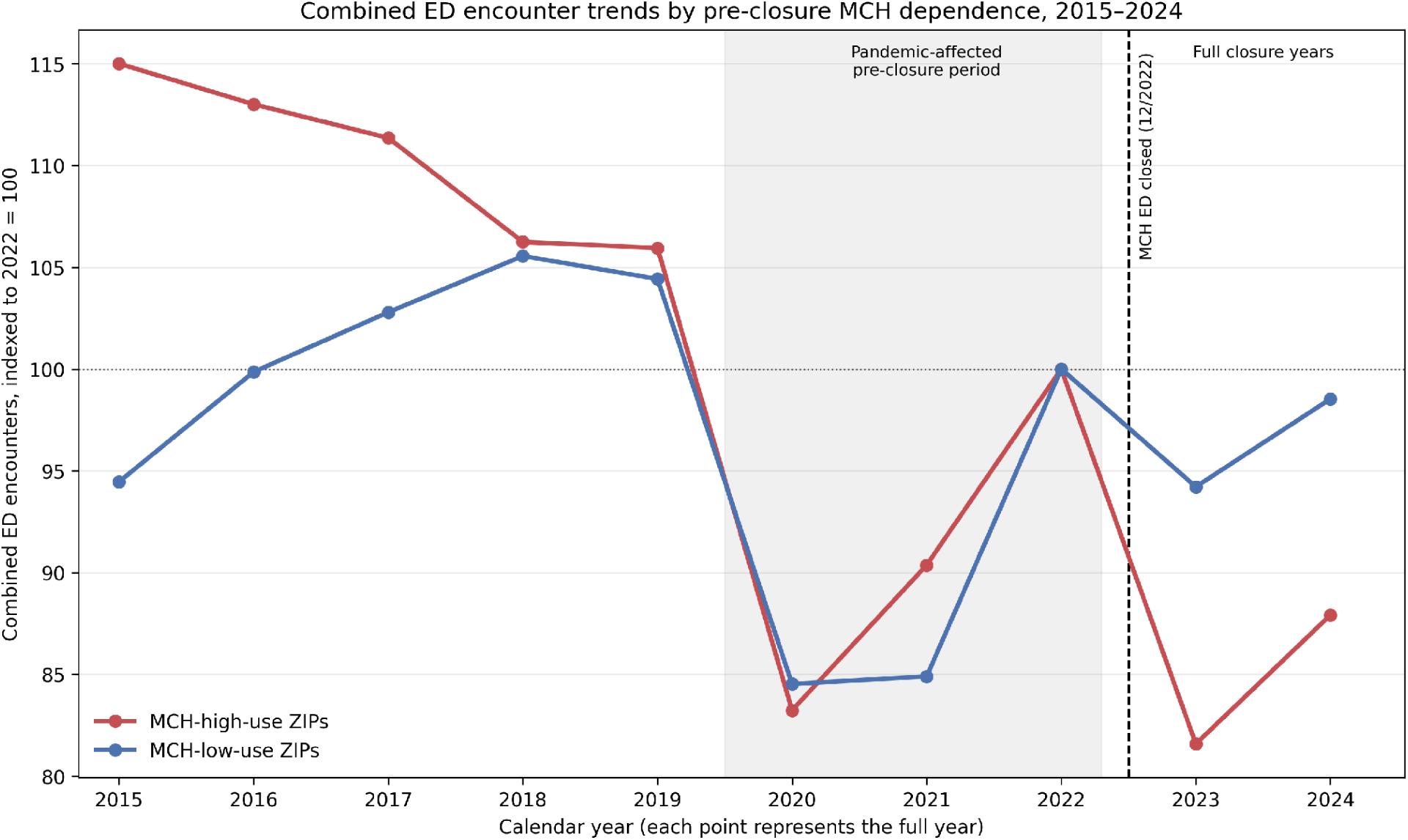
Annual combined ED encounter counts by pre-closure dependence on MCH, 2015–2024. High- and low-dependence ZIP-code groups were defined using the proportion of residents’ ED encounters occurring at MCH during 2017–2022. Each group’s annual encounter counts were indexed to its 2022 total (2022 = 100). The high-dependence group included ZIP codes 93638, 93639, 93637, and 93610; the low-dependence group included 93669, 93643, 93644, 93645, 93614, 93601, and 93604. The shaded area identifies the pandemic-affected pre-closure period (2020–2022), and the vertical dashed line marks MCH ED closure on December 30, 2022. Counts were not adjusted for population changes, and the groups had different pre-pandemic trends.

The geographic comparison was descriptive and did not adjust for annual ZIP-level population changes. In addition, the high- and low-dependence ZIP groups exhibited different pre-pandemic encounter-count trends, limiting causal interpretation of their post-closure differences. Nevertheless, the larger post-closure decline was concentrated in communities that had relied most heavily on MCH.

### Destination of emergency care

Among Madera County residents, combined ED encounters at Fresno County hospitals increased from 19,613 (31.0% of Madera-resident encounters) in 2022 to 32,878 (61.1%) in 2023. Over the same period, ED encounters among Madera County residents at hospitals located within Madera County decreased from 39,433 (62.4%) to 14,054 (26.1%), nearly all of which occurred at Valley Children’s Hospital (14,050 encounters). Encounters at Merced County hospitals increased from 1,459 (2.3%) to 3,979 (7.4%) (Table 3).

**Table 3.** Destination of combined ED encounters among Madera County residents, 2022–2023.

| Destination | 2022 encounters | 2022 share | 2023 encounters | 2023 share |
| --- | --- | --- | --- | --- |
| Madera County hospitals | 39,433 | 62.4% | 14,054 | 26.1% |
| Fresno County hospitals | 19,613 | 31.0% | 32,878 | 61.1% |
| Merced County hospitals | 1,459 | 2.3% | 3,979 | 7.4% |
| All destinations | 63,177 | 100% | 53,797 | 100% |

**Note.** Includes ED-only and inpatient-from-ED encounters among Madera County residents. Shares use annual combined ED encounters among Madera residents as denominators. Other destination counties are included in “All destinations” but are not separately displayed.

Despite this substantial redirection of care toward hospitals in neighboring counties, total combined ED encounters among Madera residents declined from 63,177 in 2022 to 53,797 in 2023. These findings document redistribution of hospital utilization but do not establish whether the overall decline reflected reduced demand, substitution to other care settings, or barriers to obtaining emergency care.

### ED-only utilization by expected payer and demographic group

The HCAI ED-only characteristics files showed decreases in Madera encounter counts across major expected-payer groups from 2022 to 2023 (Table 4): uninsured/self-pay, −30.8%; Medi-Cal, −20.9%; Medicare, −24.8%; and private insurance, −10.6%. The corresponding Fresno changes were −11.3%, +0.1%, +6.1%, and +0.1%. These represent changes in ED-only encounter counts, not utilization rates among insured or enrolled individuals. Changes in payer composition or insurance coverage may have contributed to the observed differences. The “other” category combined several heterogeneous original payer classifications.

**Table 4.**
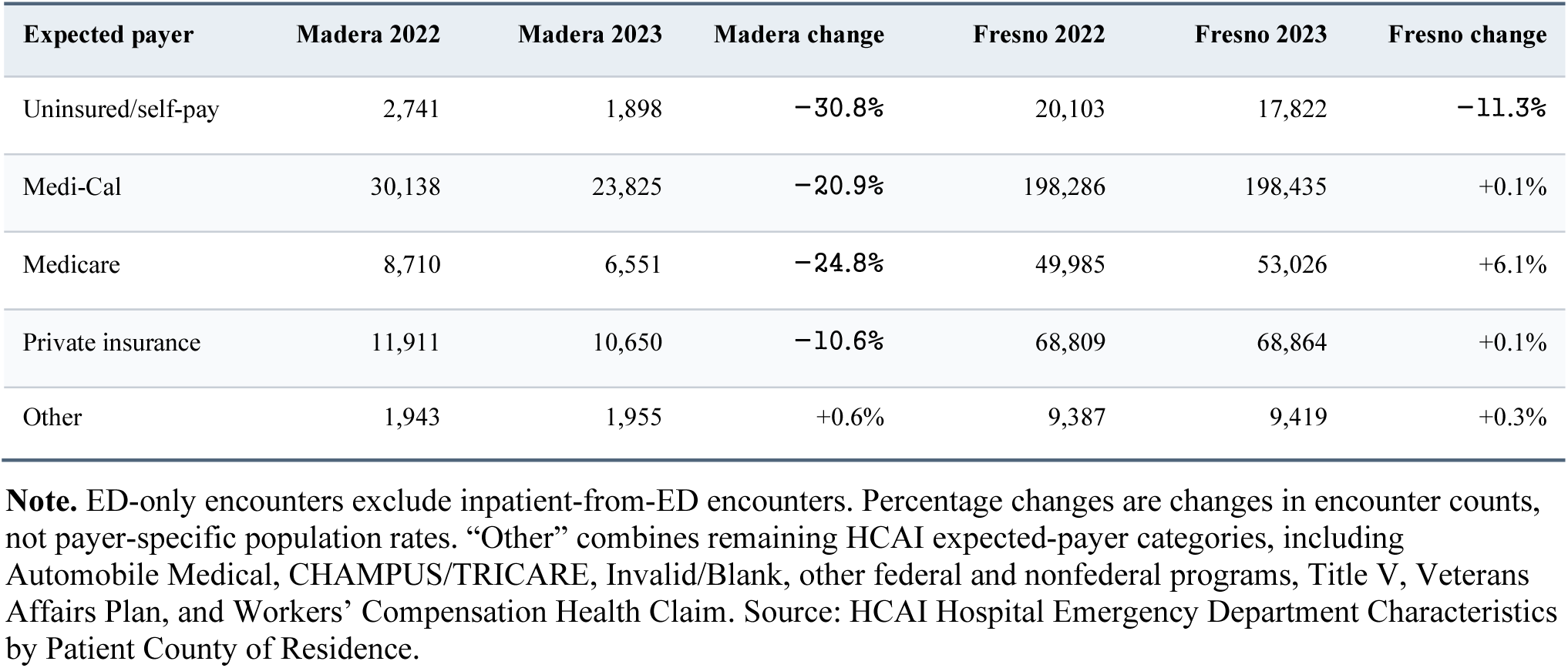
ED-only encounters by expected payer among Madera and Fresno County residents, 2022–2023.

| Expected payer | Madera 2022 | Madera 2023 | Madera change | Fresno 2022 | Fresno 2023 | Fresno change |
| --- | --- | --- | --- | --- | --- | --- |
| Uninsured/self-pay | 2,741 | 1,898 | –30.8% | 20,103 | 17,822 | –11.3% |
| Medi-Cal | 30,138 | 23,825 | –20.9% | 198,286 | 198,435 | +0.1% |
| Medicare | 8,710 | 6,551 | –24.8% | 49,985 | 53,026 | +6.1% |
| Private insurance | 11,911 | 10,650 | –10.6% | 68,809 | 68,864 | +0.1% |
| Other | 1,943 | 1,955 | +0.6% | 9,387 | 9,419 | +0.3% |

**Note.** ED-only encounters exclude inpatient-from-ED encounters. Percentage changes are changes in encounter counts, not payer-specific population rates. “Other” combines remaining HCAI expected-payer categories, including Automobile Medical, CHAMPUS/TRICARE, Invalid/Blank, other federal and nonfederal programs, Title V, Veterans Affairs Plan, and Workers’ Compensation Health Claim. Source: HCAI Hospital Emergency Department Characteristics by Patient County of Residence.

ED-only encounter counts declined across all major race/ethnicity groups examined, with larger percentage reductions among Black (30.5%) and Asian (26.4%) residents than among Hispanic (21.9%) and White (15.2%) residents. These comparisons reflect encounter counts rather than population-adjusted utilization rates, and differences in group size and reporting categories limit interpretation. Declines were similar among females (18.9%) and males (19.2%).

### Exploratory comparative interrupted time-series estimates

The primary adjusted comparative model yielded a Madera-specific closure-period combined ED rate ratio of 0.871, corresponding to an estimated 12.9% lower rate relative to the model without the Madera-specific closure interaction (Table 5). The standardized difference was approximately −51.2 combined ED encounters per 1,000 residents. Across alternative sensitivity specifications, point-estimate rate ratios ranged from 0.861 to 0.888 (approximately 11%–14% lower). The ED-only estimate was 0.848 (approximately 15.2% lower; −51.8 encounters per 1,000), with sensitivity estimates ranging from 0.831 to 0.871.

**Table 5.** Exploratory regression estimates of changes in Madera County ED utilization following MCH closure.

| Outcome | Primary rate ratio | Sensitivity range | Standardized rate difference per 1,000 residents |
| --- | --- | --- | --- |
| Combined ED | 0.871 | 0.861–0.888 | –51.2 |
| ED only | 0.848 | 0.831–0.871 | –51.8 |
| Inpatient from ED | 1.000 | 0.956–1.087 | Approximately 0 |

**Note.** Models compared Madera's 2023–2024 ED utilization with expected patterns based on Madera and Fresno County data from 2015–2024, adjusting for county-specific trends and pandemic-period changes. Rate ratios below 1.0 indicate lower Madera utilization relative to the modeled comparison. Sensitivity ranges represent estimates from alternative models, not confidence intervals. Results are exploratory because of limited post-closure observations and substantial overdispersion.

By contrast, the primary inpatient-from-ED rate ratio was approximately 1.000, with sensitivity estimates varying from 0.956 to 1.087. Thus, the descriptive increase in inpatient-from-ED utilization did not correspond to a consistent adjusted Madera-specific increase.

Because of pronounced overdispersion, the inclusion of only two counties, and the limited number of post-closure observations, model intervals and significance tests were not treated as confirmatory. The regression findings were interpreted as exploratory estimates of the association between the closure period and ED utilization, supporting but not independently establishing the patterns observed in the descriptive comparisons.

## Discussion

In this residence-based comparative study, the period following MCH closure was associated with a substantial reduction in combined ED utilization among Madera County residents relative to Fresno County residents. The decline was concentrated in ED-only encounters, while inpatient-from-ED rates increased descriptively but did not show a consistent additional Madera-specific increase in the adjusted comparative analysis. The larger decline in ED encounters in ZIP codes with high pre-closure dependence on MCH provided further descriptive evidence of geographic differences in utilization. Although the regression estimates supported the observed reduction in ED utilization, the limited number of geographic units and post-closure observations restricts causal interpretation.

### Interpretation of the ED-only decline

A hospital closure can redistribute visits to neighboring EDs without necessarily reducing total encounters among affected residents. In this study, the proportion of Madera-resident ED encounters occurring at Fresno County hospitals nearly doubled after MCH closed, yet combined utilization among Madera residents declined. Thus, increased utilization of other hospitals did not fully offset the loss of encounters occurring within Madera County. Possible explanations include increased travel time, unreliable transportation, greater cost or inconvenience, changes in healthcare-seeking behavior, reduced demand for discretionary ED services, or increased use of care settings not measured here.

The decline in ED-only utilization following MCH closure was similar in magnitude to the reduction observed during the COVID-19 pandemic. Although both ED-only and inpatient-from-ED utilization declined during the pandemic, the reduction following MCH closure was concentrated in ED-only encounters. These patterns suggest that some ED utilization may be sensitive to changes in perceived risk, accessibility, and the practical burden of obtaining care. However, reductions in utilization cannot be assumed to represent unnecessary or safely avoidable encounters, since barriers to emergency care may also reduce use for serious medical conditions.

Because the data do not link individuals or measure nonhospital services, this study cannot establish which visits were substituted, postponed, or forgone, or whether patients experienced worse outcomes.

### ED-origin admissions and regional referral networks

Before closure, Madera residents could use MCH for general emergency care, while some patients requiring specialized services were already treated at larger Fresno hospitals. These established referral patterns may have influenced the response to MCH closure. Changes after closure also may reflect hospital destination, transfer conventions, admission thresholds, and case mix. Importantly, the exploratory adjusted regression analysis did not identify a consistent additional increase in inpatient-from-ED utilization among Madera County residents relative to Fresno County residents. The observed post-closure increase in the inpatient-from-ED rate should therefore not be interpreted as evidence that MCH closure increased illness severity or delayed presentation.

### Geographic concentration and equity

The descriptive ZIP-code results suggest that the decline was concentrated in communities with the greatest pre-closure reliance on MCH. A plausible geographic explanation is that Highway 99 corridor communities had convenient access to MCH, while several Highway 41 and foothill communities historically traveled more directly to Fresno hospitals. Following MCH closure, the nearest alternative general adult emergency department was approximately 23 miles away by highway, although the additional travel distance varied according to patients’ locations of residence. Valley Children’s Hospital remained available for pediatric emergency care within Madera County but did not replace the general adult emergency services previously provided by MCH. The resulting increase in travel distance may have posed particular difficulties for residents with limited transportation resources.

For households with limited access to transportation, an increase in travel time may be more consequential than for households with reliable vehicles. These are hypotheses rather than measured mechanisms; driving times, vehicle availability and confounding by socioeconomic differences among ZIP codes were not directly analyzed in this manuscript. The low-dependence comparison was also not an untreated control: regional system congestion or referral changes could have affected those communities.

Because the high- and low-dependence ZIP-code groups differ in geography, demographics, economic conditions, and historical ED trajectories, the larger decline in the high-dependence group should be viewed as supporting descriptive evidence rather than a separate controlled natural experiment. The county comparison remains the primary population-adjusted analysis, while the ZIP-code comparison helps characterize heterogeneity within Madera County.

The magnitude of the decline in ED-only encounters among uninsured and publicly insured patients is particularly noteworthy. From 2022 to 2023, encounters declined by 30.8% among uninsured/self-pay patients, 24.8% among Medicare patients, and 20.9% among Medi-Cal patients, compared with 10.6% among privately insured patients. These differences raise concerns that the loss of a local ED may have disproportionately affected populations facing barriers to alternative emergency care. Because these comparisons reflect encounter counts rather than population-adjusted rates, changes in insurance enrollment or patient characteristics may have contributed to the observed patterns. The larger declines in ED-only encounters among Black (30.5%) and Asian (26.4%) residents, compared with White residents (15.2%), also raise questions about potential disparities in access to emergency care following hospital closure. Differences in population size, demographic composition, and potential inconsistencies in race/ethnicity reporting limit interpretation. These findings warrant further investigation but do not establish differential effects of the closure across racial or ethnic groups.

### Relation to prior literature and implications

Previous studies of hospital closures have documented increased ED volume at neighboring hospitals, changes in regional emergency care capacity, and potential consequences for patient outcomes. Lee and colleagues, analyzing hospital closures in New York State, found that proximity to a hospital closure was associated with increased ED volume at surrounding hospitals.^15^ A study of rural hospital closures similarly documented increased ED visits and inpatient admissions at neighboring hospitals.^5^ These studies demonstrate how closures can redistribute utilization among healthcare institutions, but do not establish whether total ED utilization among residents of affected communities remains unchanged.

Other research has examined utilization among residents of affected communities. A study of commercially insured individuals in Texas found reductions in ED and outpatient utilization following rural hospital closures.^6^ These findings suggest that hospital closure may be associated with reduced healthcare utilization even when alternative facilities remain available. The present study extends this literature by examining population-based ED utilization across payer groups, distinguishing ED-only from inpatient-from-ED encounters, and characterizing geographic differences according to preclosure reliance on MCH.

The findings here support planning for transportation and access to emergency care when local hospital services close, but they do not establish whether reduced utilization represents changes in demand, substitution to other services, or unmet medical need. The study also does not determine the optimal alternative service model or quantify downstream health or financial consequences.

### Strengths and limitations

Strengths include a validated statewide administrative source; analysis by residence rather than facility location; separate measurement of ED-only and inpatient-from-ED encounters; official annual county populations; pre-closure ZIP-code reliance measures; alternative comparative model specifications; and a documented independent reproduction audit. Nevertheless, the observational comparison involved only two counties and two complete closure years. Both counties experienced major pandemic-related utilization disruptions, underlying pre-pandemic trends were not perfectly parallel, and Fresno hospitals may have experienced spillovers from displaced Madera patients. The rate model had substantial overdispersion and too few independent geographic units for reliable conventional confidence intervals. Sensitivity agreement does not make the underlying counterfactual assumption verifiable.

Additional limitations include annual rather than monthly observations, different county population-estimate vintages over the study window, inability to link patients or transfers across facilities, and possible changes in administrative classification or reporting. ZIP-code comparisons used postal ZIP-code encounter counts without reliable annual ZIP-code denominators; preexisting trends and population changes complicate their causal interpretation. The study did not measure acuity, potential avoidability, outpatient substitution, transport time, mortality, unmet care, or cost. These questions should be evaluated separately rather than inferred from the observed utilization trends. In addition, although the public HCAI encounter totals were independently reproduced, the documentation did not fully establish whether small-cell suppression affected the ZIP-code-level source data.

HCAI expected-payer, race/ethnicity, and sex files measure ED-only encounters and cannot directly explain the combined ED outcome or infer risk at the individual level. Their published category structures and potential administrative coding changes add limitations to subgroup interpretation.

## Conclusions

Population-adjusted ED utilization fell substantially among Madera County residents during the two full years following MCH closure, relative to Fresno County residents and modeled prior trends. The reduction was concentrated in ED-only encounters and geographically more pronounced in communities with high pre-closure reliance on MCH. Although comparative point estimates were consistent across specifications, the limited series and inferential constraints preclude definitive causal attribution. Further work is needed to establish whether reduced ED use reflected substitution, changes in demand, or barriers to necessary care.

## Data Availability

https://data.chhs.ca.gov/dataset/patient-origin-market-share-pivot-profile-inpatient-emergency-department-and-ambulatory-surgery

https://data.chhs.ca.gov/dataset/hospital-emergency-department-characteristics-by-patient-county-of-residence

https://dof.ca.gov/Forecasting/Demographics/Estimates/

https://dof.ca.gov/Forecasting/Demographics/Estimates/E-2/

